# Contemporary social context and patterns of prenatal cannabis use in Canada following legalization: a secondary analysis of prospective cohort data

**DOI:** 10.1101/2022.06.20.22276670

**Authors:** Kathleen H. Chaput, Harleen Sanghera, Sanam Sekandary, Carly McMorris, Amy Metcalfe, Stephen Wood, Deborah McNeil, Sheila McDonald

## Abstract

**Background:** The epidemiology of prenatal cannabis use in Canada following legalization remains unknown despite increasing evidence for associated health risks. Our study aimed to identify current risk factors for, and patterns of, prenatal cannabis use and second-hand cannabis exposure in Alberta.

**Methods:** We conducted a secondary analysis of prospective data from a 2019 study in Calgary AB, of 153 pregnant (<28 weeks gestation at enrollment), English-speaking Alberta residents. We conducted descriptive analyses of prenatal cannabis use patterns (timing, frequency, dose, modes and reasons for use) and logistic regression to identify risk factors for direct use and second-hand exposure.

**Results:** Odds of prenatal cannabis use were significantly higher among those who did not own their home (Odds Ratio (OR) 3.1; 95% CI:1.6-9.6), smoked tobacco prenatally (OR 3.3,95% CI:1.2-9.3) and used illicit substances in the past (OR 3.2; 95% CI:1.7-9.9), and lower for those consuming alcohol prenatally (OR 0.3, 0.12-0.89). Among the 90 (58%) participants who used cannabis prenatally, the majority used for medicinal reasons (96%), at least daily (67%), by smoking (88%), in all trimesters of pregnancy (66%). Although reported dose-per-use was commonly low, cumulative doses over pregnancy were high.

**Interpretation:** Our study finds marked differences in prenatal cannabis use risk factors, and patterns of more frequent use sustained throughout pregnancy with perceived medicinal indications than pre-legalization studies. Prenatal care providers should include cannabis explicitly in medication counselling. Further prospective studies are needed as the impacts of prenatal cannabis on maternal and infant health in Canada may currently be underestimated.

## Introduction

Following cannabis legalization in Canada(1) concern about prenatal cannabis use is increasing among health professionals, and Canadian and U.S. governments have identified it as a health research priority.(1–3) Studies of prenatal cannabis use and adverse maternal and child health outcomes have found associations with low birth weight, small for gestational age, placental abruption, preterm delivery, stillbirth, maternal anemia, low APGAR scores, Neonatal Intensive Care Unit admission, and child neurodevelopment(2,8-12), though findings are mixed, due partly to the inability to control for high degrees of poly-substance use.(4,5)

Previous studies of risk factors for prenatal cannabis use include low education, young age, low income, lone-parent household, non-white ethnicity, and concurrent alcohol, tobacco, and other substance use.(6–9) in addition to mental health issues, and abnormal body mass index.(10,11) While a few Canadian studies of risk factors for prenatal cannabis use exist, (10– 12) all were conducted prior to legalization, and may be hindered by outdated data(12) and risk of misclassification bias from measurement of prenatal cannabis use with administrative data.(10,11) Relying on administrative data measures is problematic due to known high rates of underreporting substance use (60%-80%) to care providers(13), and lack of systematic screening throughout pregnancy.(14,15) Previous studies may thus not accurately reflect risk factors for use in the current Canadian context.

The few studies that have examined patterns of prenatal cannabis use found use is most common in the first trimester tapering off thereafter(6,16–18), that it’s most commonly smoked(19), and that use is largely recreational.(6) However, with trends of increasing use of cannabis for medicinal purposes, the proliferation of vape, edible/oral, and topical products available, along with improving social acceptability of cannabis use observed to accompany legalization(20,21), the extent of cannabis use throughout pregnancy may be underestimated. Further, evidence regarding the prevalence of second-hand cannabis exposure during pregnancy and whether it is associated with health outcomes is lacking.

Objective: to identify the risk factors for, and patterns of cannabis use among pregnant Canadians post-legalization.

## Methods

We conducted a secondary analysis of data from the prospective Cannabis Exposure in Pregnancy Tool (CEPT) Study.(22) The CEPT study recruited participants from July-December 2019, using: 1) recruitment letters mailed to patients who visited any Alberta Health Services (AHS) clinic for pregnancy-related care in the preceding six months, identified using pregnancy-related codes in the National Ambulatory Care Reporting System (NACRS) (Appendix A), and social media advertising targeted to women aged 18-45, residing in Alberta, with interests in pregnancy, parenting, and/or cannabis, as well as gender-neutral recruitment ads posted in transgender groups. Research assistants screened interested participants for eligibility by phone, and obtained written informed consent. We included those who were currently pregnant, and ≥18 years old, residing in Alberta, Canada. We excluded participants who were beyond 28 weeks’ gestation, to allow completion of data collection prior to delivery, and those who were not pregnant, or unable to complete English questionnaires. The University of Calgary Conjoint Health Research Ethics Board approved this Study (Ethics ID: REB19-0670).

The CEPT study included identical electronic questionnaires at enrollment and 3-month follow-up. We analyzed data from all participants with 3-month follow-up data (N=153). Any missing data on the follow-up questionnaire were imputed using enrollment questionnaire data, and any remaining missing data were handled by pairwise deletion. We measured prenatal cannabis exposure using the standardized CEPT, which measures frequency, estimated dose, timing (trimester) of use, reasons for, and forms of consumption, as well as second-hand exposure, and has excellent discriminant (Cohen’s kappa=-0.27-0.15) and convergent (Cohen’s kappa=0.73-1.0) validity, internal consistency (Cronbach’s alpha=0.92), and test-retest reliability (weighted Kappa=0.92, 95% CI:0.86 – 0.97).(22) The questionnaire also included measures of tobacco and alcohol consumption, partner, family and friends’ substance use history, self-reported mental and physical health, parity, and socioeconomic variables.

We summarized participant characteristics and patterns of prenatal cannabis consumption including any prenatal cannabis use, reasons for use and product type, timing, frequency, and estimated doses of exposure using descriptive statistics. We derived dose over pregnancy using self-reported dose-per-use, multiplied by frequency of use and trimesters in which use was reported. We assigned a conservative daily frequency of 2 consumptions for each day of “multiple times per day” use reported, and used the highest category of dose-per-use if different doses were reported for multiple cannabis products. We conducted a Chi-square test for difference in proportions to determine whether dose-per-use differed by frequency of use. We also calculated the frequency and timing in pregnancy of exposure to second-hand cannabis smoke and/or vapour while in the same room as the person using. We conducted multivariable logistic regression modelling to identify risk factors for prenatal cannabis use and second-hand exposure. We selected potential risk factors a-priori based on previous literature and used a rule of thumb of a minimum of 8 outcomes per explanatory variable included in the model.(23) Potential risk factors were household income (above or below $60,000 annually), maternal education (postsecondary/high-school or less), marital status (married or common-law vs. other), visible minority (Y/N), housing ownership (Y/N), urban vs. rural residence, self-rated mental health, alcohol and tobacco use in pregnancy (Y/N), history of heroin or cocaine use (Y/N), partners’ and close friends’ having substance use problems (Y/N). We used Stata, version 17BE, to conduct all analyses at a two-tailed alpha-level of 0.05.

## Results

Participant characteristics are reported in Table 1 and were generally similar to population averages in the province of Alberta (Figure 1). In the full sample of 153 pregnant Albertans, 90 participants (58.8%) reported cannabis consumption at least once during pregnancy. Multivariable logistic regression revealed significantly higher odds of prenatal cannabis use among those who did not own their home (adjusted odds ratio (aOR) 3.1, p=0.02), smoked tobacco in pregnancy (aOR 3.3, p=0.02) and had used illicit substances in the past (aOR 3.2, p=0.04), and significantly lower odds among those who consumed alcohol prenatally (aOR 0.3, p=0.03). (Table 2) We found significantly higher odds of second-hand cannabis exposure for those who did not own their homes (aOR 6.6, p<0.001), and had friends with substance use concerns (aOR 2.2, p=0.049) (Table 3).

**Table 1:** Participant Characteristics (Variable-specific *n* differs due to pairwise deletion of missing data)

| Variable |  | Consumed Cannabis | Total Sample |
| --- | --- | --- | --- |
|  |  | *n* (%)<br>90 (58.8) | *n* (%)<br>153 (100) |
| **Parity** |  |  |  |
|  | 0 | 33 (39.3) | 54 (36.7) |
|  | >1 | 51 (60.7) | 93 (63.3) |
| **Marital Status** |  |  |  |
|  | *Married/Common Law* | 61 (69.3) | 117 (77.5) |
|  | *Other* | 27 (30.7) | 34 (22.5) |
| **Household annual income** |  |  |  |
| | < \$60,000 | 41 (47.7) | 60 (40.3) |
| | \$60,000 or more | 45 (52.3) | 89 (59.7) |
| **Home ownership** |  |  |  |
|  | *Owns home* | 29 (33.3) | 75 (50.0) |
|  | *Non-owner* | 58 (66.7) | 75 (50.0) |
| **Area of residence** |  |  |  |
|  | *Urban* | 67 (75.3) | 115 (75.7) |
|  | *Rural* | 22 (24.7) | 37 (24.3) |
| **Minority Status** |  |  |  |
|  | *Visible minority* | 16 (17.8) | 23 (15.7) |
|  | *Not a visible minority* | 74 (82.2) | 124 (84.3) |
| **Education** |  |  |  |
|  | *High School or less* | 40 (46.5) | 18 (12.1) |
|  | *Some post-secondary* | 46 (53.5) | 31 (20.8) |
| **Self-rated mental health** |  |  |  |
|  | *Good/V. good/Excellent* | 68 (79.1) | 125 (83.9) |
|  | *Poor/Terrible* | 18 (20.9) | 24 (16.1) |
| **Alcohol in pregnancy (any)** |  |  |  |
|  | *Yes* | 20 (23.8) | 42 (28.6) |
|  | *No* | 64 (76.2) | 105 (71.4) |
| **Tobacco in pregnancy (any)** |  |  |  |
|  | *Yes* | 39 (45.9) | 48 (32.4) |
|  | *No* | 46 (54.1) | 100 (67.6) |
| **Friends with substance concerns** |  |  |  |
|  | *Yes* | 42 (48.8) | 78 (55.3) |
|  | *No* | 40 (51.2) | 63 (44.7) |
| **Partner with substance concerns** |  |  |  |
|  | *Yes* | 15 (18.5) | 18 (12.6) |
|  | *No* | 66 (81.5) | 125 (87.4) |
| **History of heroin/cocaine use (ever)** |  |  |  |
|  | *Yes* | 31 (42.5) | 42 (28.6) |
|  | *No* | 42 (57.5) | 105 (71.4) |
| **Second-hand cannabis smoke/vapour exposure** |  |  |  |
|  | *Yes* | 71 (78.9) | 85 (55.6) |
|  | *No* | 19 (21.1) | 68 (44.4) |
| **Maternal age (years)\*** |  | -- | **Mean (SD)**<br>27.2 (7.6) |

\*aggregate measure

**Table 2:**
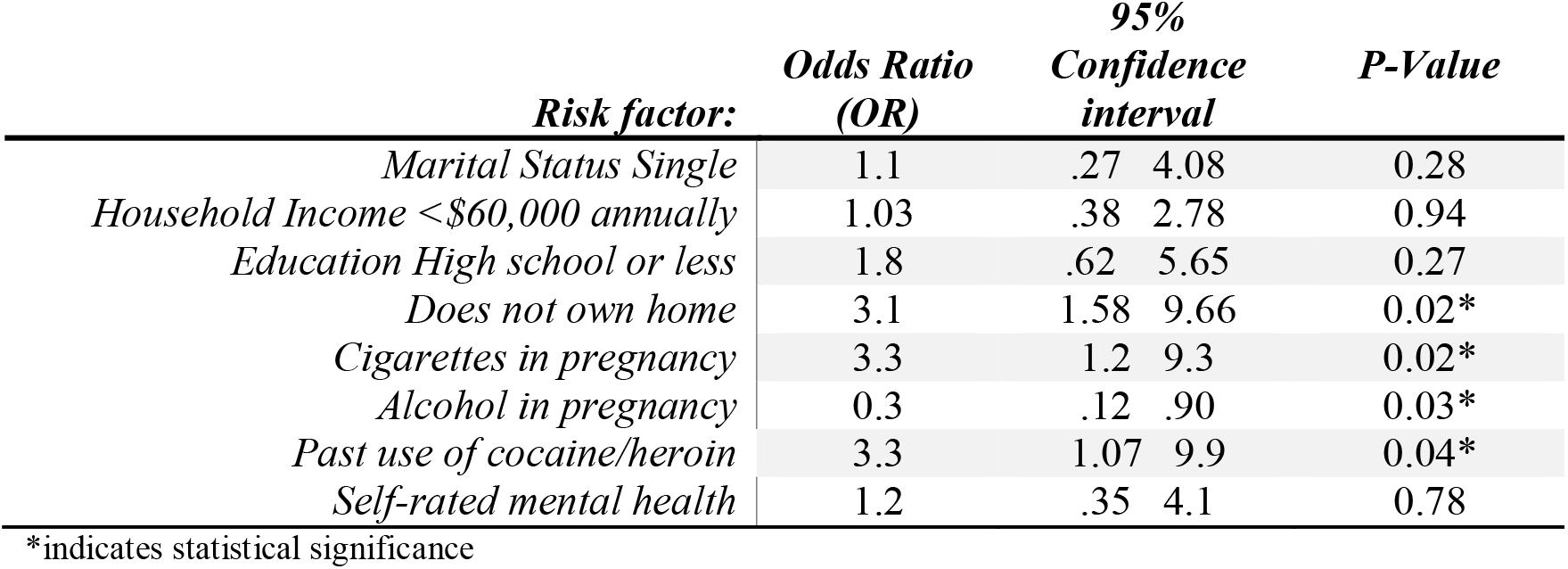
Risk factors for any prenatal cannabis use

**Table 3:**
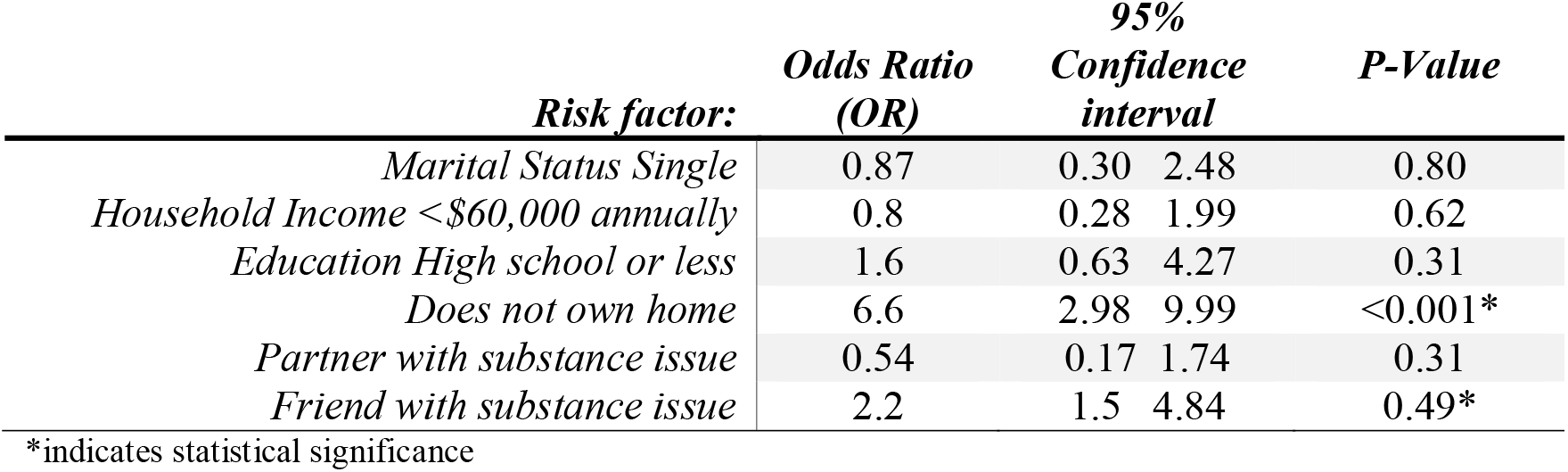
Risk factors for second-hand exposure to cannabis smoke or vapour

| ***Risk factor:*** | ***Odds Ratio***<br>***(OR)*** | ***95%***<br>***Confidence***<br>***interval*** |  | ***P-Value*** |
| --- | --- | --- | --- | --- |
| *Marital Status Single* | 0.87 | 0.30 | 2.48 | 0.80 |
| *Household Income <\$60,000 annually* | 0.8 | 0.28 | 1.99 | 0.62 |
| *Education High school or less* | 1.6 | 0.63 | 4.27 | 0.31 |
| *Does not own home* | 6.6 | 2.98 | 9.99 | <0.001* |
| *Partner with substance issue* | 0.54 | 0.17 | 1.74 | 0.31 |
| *Friend with substance issue* | 2.2 | 1.5 | 4.84 | 0.49* |

\*indicates statistical significance

**Figure 1:**
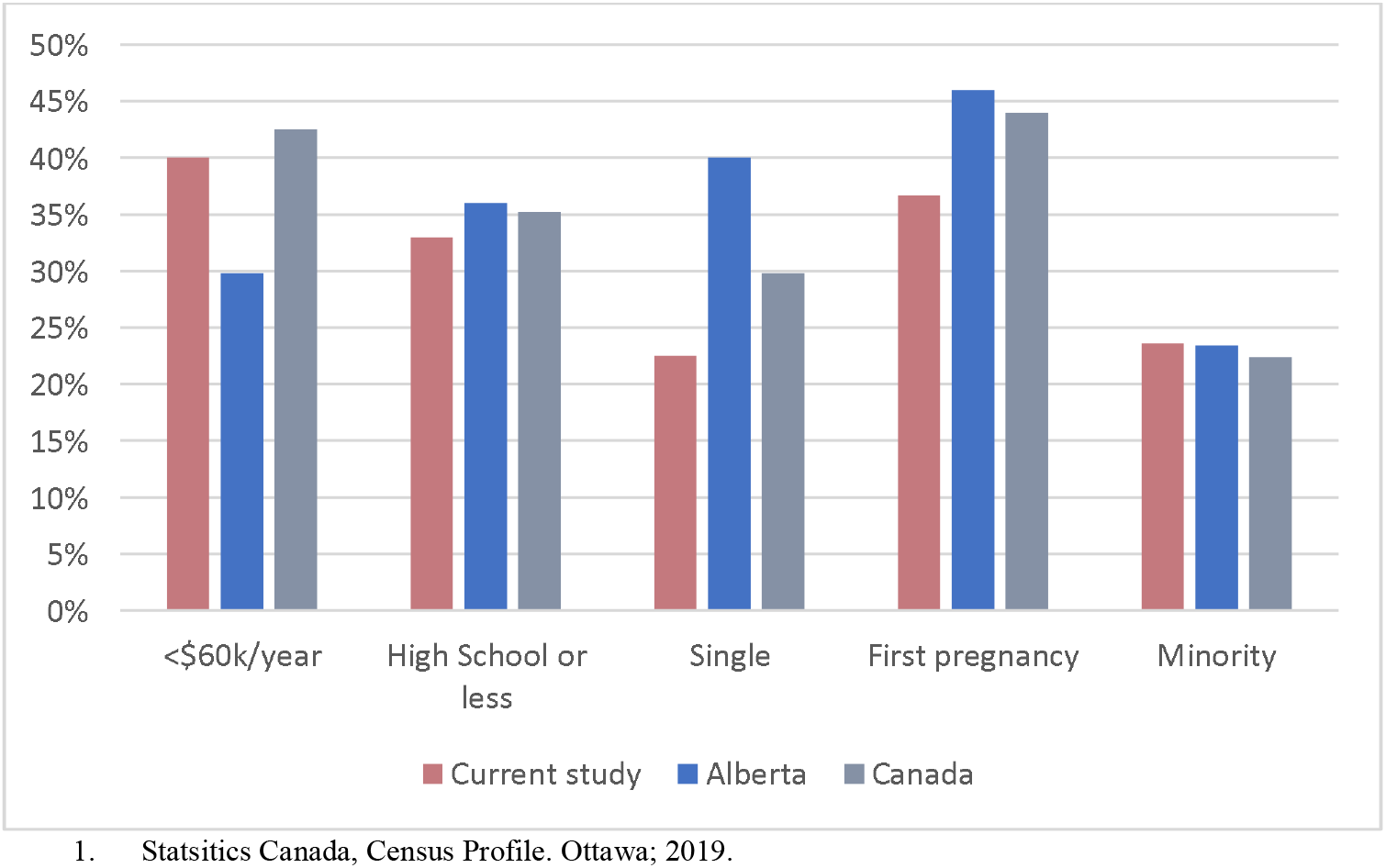
Participant characteristics compared to population of Alberta and Canada^1^

Patterns of prenatal cannabis consumption appear in Table 4. Nearly all (95.6%) of those who used cannabis reported at least one perceived medicinal reason for use. The most reported mode of cannabis consumption was smoking (88.4%), followed by concentrated products (20.9%), with 25% of consumers reporting use of CBD-only products. Controlling for weeks of gestation at survey completion, cannabis use was highest in the first trimester (86%). No significant change in proportion of use in the second (74%) and third (81%) trimesters was observed (p=0.79), and most cannabis consumers (66%) reported use in all trimesters. Use ranged in frequency from “once or twice, but less than monthly” (11.8%) to multiple times per day (15%), with 68.7% of users reporting at least daily consumption. Forty percent of consumers reported very low dose-per-use (<10mg THC inhaled, <15mg oral), and 28% high or very high dose-per-use (60 to >240mg THC). Total dose over pregnancy varied, with cumulative dose increasing exponentially with increasing frequency of use (Figure 2). A higher proportion of those using at least daily reported moderate to very high dose-per-use (20.7% vs. 54.5%; p=0.003).

**Table 4:** Patterns of Cannabis Use in Pregnancy

| Variable | Cannabis consumers<br>N (%) | Full Sample<br>N (%) |
| --- | --- | --- |
| **Reason for use<sup>a</sup>** |  |  |
| *Nausea and vomiting* | 72 (80.0) | -- |
| *Sleep* | 58 (64.4) | -- |
| *Anxiety* | 54 (60.0) | -- |
| *Appetite* | 52 (57.8) | -- |
| *Pain* | 44 (48.9) | -- |
| *Depression* | 35 (38.9) | -- |
| *Recreation* | 57 (63.3) | -- |
| **Form of cannabis consumption<sup>a</sup>** |  |  |
| *Smoke* | 79 (87.8) | -- |
| *Vape* | 10 (11.1) | -- |
| *Oral/Edible (with THC)* | 14 (15.6) | -- |
| *Topical (with THC)* | 4 (4.4) | -- |
| *Concentrate* | 19 (21.1) | -- |
| *Oral/Edible (CBD-only)* | 14 (15.6) | -- |
| *Topical (CBD-only)* | 8 (8.9) | -- |
| **Timing of use** |  |  |
| *First Trimester (n=90)* | 78 (86.7) | -- |
| *Second Trimester (n=84)* | 63 (75.0) | -- |
| *Third Trimester(n=70)* | 58 (84.5) | -- |
| *All Trimesters<sup>b</sup>* | 59 (65.6) | -- |
| **Frequency of use** |  |  |
| *Once or twice, less than monthly* | 18 (20.2) | -- |
| *Monthly* | 7 (7.9) | -- |
| *Weekly* | 9 (10.1) | -- |
| *Daily* | 32 (36.0) | -- |
| *Multiple times per day* | 23 (25.8) | -- |
| **Dose-per-use** |  |  |
| *Low* | 36 (40.0) | -- |
| *Medium* | 17 (18.9) | -- |
| *Moderate* | 12 (13.3) | -- |
| *High* | 9 (10.0) | -- |
| *Very High* | 16 (17.8) | -- |
| **Second-hand Frequency** |  |  |
| *<Monthly* | 12 (16.9) | 23 (27.1) |
| *Monthly* | 7 (9.9) | 9 (10.6) |
| *Weekly* | 11 (15.5) | 11 (12.9) |
| *Daily* | 26 (36.6) | 27 (31.8) |
| *Multiple times per day* | 15 (21.1) | 15 (17.7) |
| **Second-hand timing** |  |  |
| *First Trimester* | 79 (93.0) | 72 (84.7) |
| *Second Trimester* | 60 (76.0) | 65 (76.5) |
| *Third Trimester* | 60 (76.0) | 38 (44.7) |

a More than one choice permitted; b controlling for gestational age

**Figure 2:**
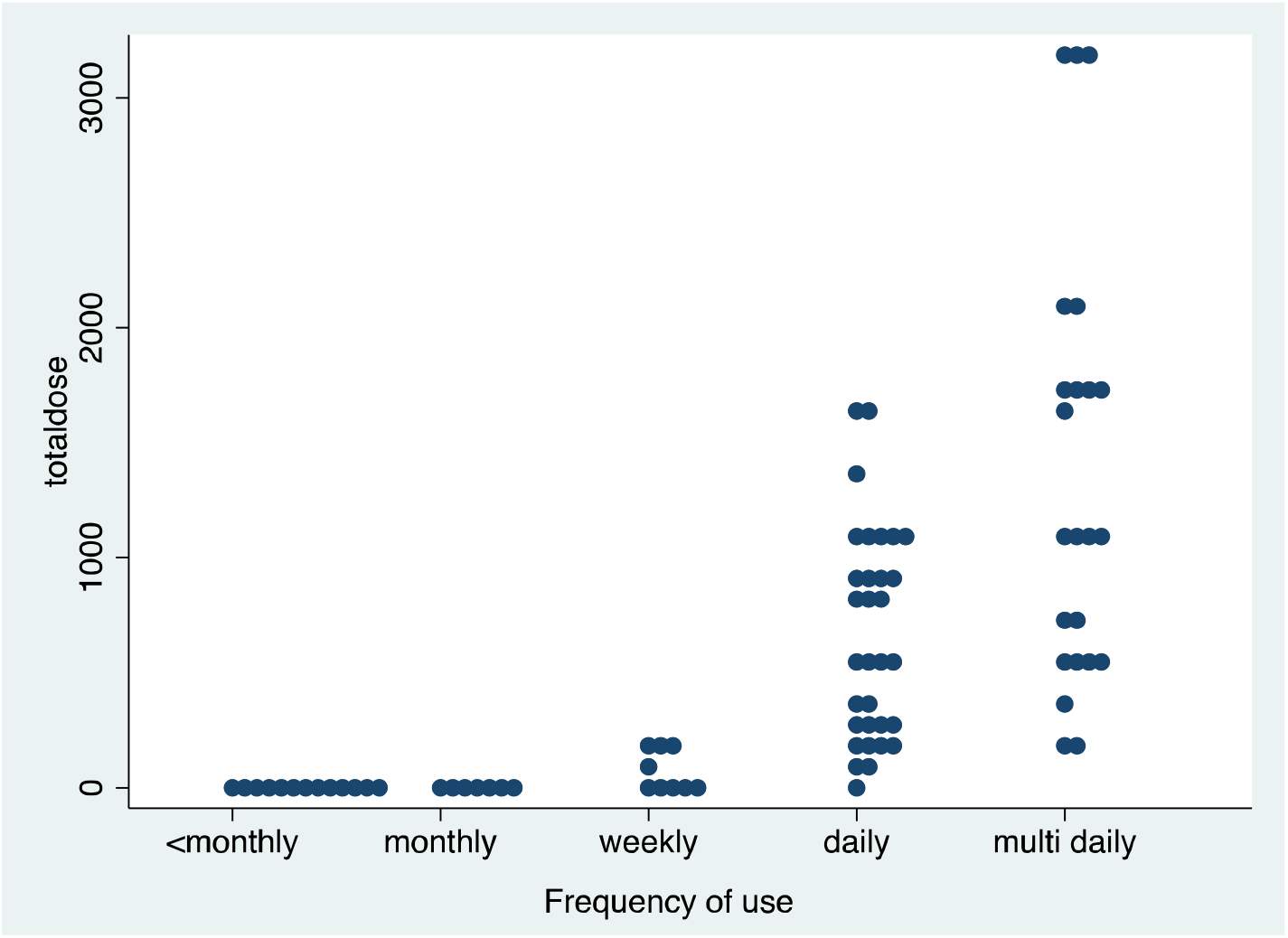
Cumulative dose (total dose) over pregnancy by frequency of use

Fifty-six percent of all participants and 79% of those who used cannabis were exposed to second-hand cannabis smoke or vapour, and of those, half (49.4%) were exposed at least daily, with exposure most frequently occurring in the first trimester (93%) and the majority (73%) having sustained exposure throughout all trimesters.

## Interpretation

In our secondary analysis of prospective post-legalization study data, 59% of participants (n=90) consumed cannabis at least once during pregnancy, reflecting the targeted over-recruitment of cannabis users to the original study, rather than a true prevalence of prenatal cannabis use in Alberta. Our study found that the odds of prenatal cannabis use were significantly higher for those who did not own their homes, concurrently smoked and had a history of using other illicit substances, but lower for those who drank alcohol prenatally, and did not differ with other sociodemographic factors. We found that almost all prenatal cannabis consumers perceived medicinal reasons for use, highlighting a need for prenatal care providers to include cannabis explicitly in discussions of symptom management and medication use during pregnancy, to ensure appropriate prenatal counselling and informed decision making. Most prenatal consumers use cannabis at-least daily, and sustain use throughout pregnancy, leading to high cumulative doses of THC exposure.(Figure 2) Exposure to second-hand smoke and vapour is also common and may be an important route of exposure in pregnancy.

The risk factors for prenatal cannabis use identified in our study contrast with those in previous studies that reflect lower socioeconomic status (low income, single, low education), non-white ethnicity, anxiety, and use of other substances prenatally including alcohol.(6,10–12,18,24) While non-homeownership was significantly related to cannabis use in our study, our risk factor analysis supports a difference in the characteristics of cannabis users post-legalization in Alberta. Most cannabis consumers had high household incomes, post-secondary education, were in stable partnerships, did not identify as minorities, and had high self-rated mental health, supporting that universal screening may be useful. Notably, we found a decreased odds of prenatal cannabis use among those who drink alcohol in pregnancy. While this finding differs from previous research(6,7,11,12,18,24,25), it is congruent with recent qualitative research on attitudes and beliefs about prenatal cannabis use in Alberta, in which cannabis was seen as safer in pregnancy relative to other substances, including prescription medications and alcohol.(26) This finding also aligns with the majority of our sample reporting medicinal reasons for use, in contrast to predominantly recreational use reported previously.(6) The significantly higher odds of cannabis consumption with concurrent tobacco use and history of other illicit substances were congruent with previous studies.(6,7,11,12) Our findings that second-hand cannabis exposure is prevalent particularly among those with friends who have substance concerns indicates that social situations could be a significant source of exposure and pregnant people should be made aware. Although data on the impact of second-hand cannabis exposure on maternal and infant health is limited, research suggests that blood-THC levels of those exposed to second-hand cannabis smoke in a shared room can reach levels equivalent to the active user’s within 20 minutes.(27) Second-hand exposure is thus a possible mechanism for previously reported links between partner’s cannabis use and adverse infant outcomes.(28,29)

The patterns of prenatal cannabis use revealed in our analyses fill an important knowledge gap. The high prevalence of medicinal reasons for use is a novel finding that may reflect a perception of lower stigma around medicinal use of cannabis post-legalization.(20) It is also congruent with qualitative evidence that cannabis is perceived as highly effective for multiple conditions in pregnancy including nausea and vomiting, anxiety, sleep, and pain.(30,31) This highlights a need for prenatal care providers to include cannabis explicitly in discussions about safe, effective medication use, recognizing that a lack of symptom control may lead to self-medication. Our finding of predominant consumption by smoking, aligns with other prenatal studies(6) as well as data for the general Canadian population(21), however, a higher prevalence of concentrated product use in our study compared to previous research and the general population is concerning.

Recent qualitative studies indicate that during pregnancy people often reduce the amount of cannabis consumed per use, but increase their frequency of use, particularly for control of nausea and to help with appetite.(26,31) While this aligns with the high prevalence of daily or more frequent consumption in our study, we found that who use cannabis more frequently more commonly report higher dose-per-use. Further, the majority of consumers sustained use throughout pregnancy, contrasting previous studies that found sharp drops in use after the first trimester. This is particularly concerning in light of the evidence that the potency of cannabis on the North American market has drastically increased in recent years(32,33), and supports that previous studies may be underrepresenting the impact of prenatal cannabis use on the health outcomes of Canadian children.

Our study findings collectively outline different patterns of use and a different social context for prenatal cannabis use in Canada, currently, compared with previously published literature. Pre-legalization and non-Canadian studies may be supporting assumptions about how, why, and by whom cannabis is consumed in pregnancy, that could be hindering adequate and effective public health action and clinical screening and counselling. Further, there is an urgent need for high-quality prospective studies that capture the large cumulative doses of cannabis sustained throughout pregnancy observed in our study, to validly capture the effects of prenatal cannabis exposure on maternal and infant health.(1-3) Overlapping alcohol, tobacco and cannabis use was lower than in previous studies, indicating their confounding and potentially modifying effects can be properly assessed in future research, compared to older studies that were underpowered to separate effects of large overlap.(2) There is currently a lack of evidence on the relative safety of smoking versus other forms of prenatal cannabis consumption which should be examined in future studies. Nearly a quarter of cannabis consumers reported using CBD-only products prenatally, and future studies should examine whether maternal and infant health outcomes are associated with prenatal exposure to CBD-only products and those containing THC separately.

Strengths and limitations: Our findings must be interpreted within the context of our study’s limitations. The relatively small sample size of our study limited our ability to examine a broader set of socioeconomic risk factors for cannabis use, as the prevalence of minority status and rural residence in the non-user group was very low, precluding us from including these variables in our models. However, the over-recruitment of cannabis consumers did increase power, allowing us to examine important risk factors for use, and our sample was generally representative of maternal populations in Alberta and Canada(34) (Figure 1). A further limitation of this study was our lack of an individual measure of maternal age, which should be examined as a potential risk factor in future studies. Lastly, while we attempted to recruit gender-diverse participants, our study sample was all female-identifying. Future research should examine the patterns of prenatal cannabis exposure using a larger and more diverse samples to confirm our findings. A major strength of our study was the use of a detailed standardized measure of prenatal cannabis use, developed and validated for pregnant populations.(22)

Our results reveal marked differences between the current, post-legalization patterns of prenatal cannabis consumption in Alberta, and previously published evidence. Improved understanding of the patterns of prenatal cannabis consumption and second-hand cannabis exposure in the current Canadian context is essential to our understanding of the maternal and infant health impacts of prenatal cannabis use in the Canadian population. Our study can help to inform properly targeted public health measures and prenatal care. Given the current legal availability of myriad cannabis products, the drastic increases in cannabis potency, and rising prenatal cannabis use since legalization(20), our findings of predominant high-frequency use throughout pregnancy, indicate that the current evidence may be underestimating the actual maternal and infant health impacts of prenatal cannabis occurring in Canada. Our study further highlights urgent need for prospective research with high quality measurement, to improve the evidence base and allow for adequate public health education, informed clinical care, and ultimately improved maternal and infant health.

## Data Availability

The datasets generated and analysed during the current study are not publicly available due to privacy restrictions outlined in the ethics agreement. De-identified may be made available from the corresponding author on reasonable request following publication.

## Notes

<u>Author conflict statements:</u> No Authors have any conflicts of interest to declare.

### Competing Interest Statement

The authors have declared no competing interest.

### Funding Statement

This project was funded by a research grant from the Alberta Childrens Hospital Research Institute.

### Author Declarations

The University of Calgary Conjoint Health Research Ethics Board approved this Study (Ethics ID: REB19-0670).

